# Millimeter-sized battery-free epidural cortical stimulators

**DOI:** 10.1101/2023.09.13.23295460

**Authors:** Joshua E. Woods, Amanda L. Singer, Fatima Alrashdan, Wendy Tan, Chunfeng Tan, Sunil A. Sheth, Sameer A. Sheth, Jacob T. Robinson

## Abstract

Refractory neurological and psychiatric disorders are increasingly treated with brain stimulation therapies using implanted neuromodulation devices. Current commercially available stimulation systems, however, are limited by the need for implantable pulse generators and wired power; the complexity of this architecture creates multiple failure points including lead fractures, migration, and infection. Enabling less invasive approaches could increase access to these therapies. Here we demonstrate the first millimeter-sized leadless brain stimulator in large animal and human subjects. This Digitally programmable Over-brain Therapeutic (or DOT) is approximately 1 cm in width yet can produce sufficient energy to stimulate cortical activity on-demand through the dura. This extreme miniaturization is possible using recently developed magnetoelectric wireless power transfer that allows us to reach power levels required to stimulate the surface of the brain without direct contact to the cortical surface. This externally powered cortical stimulation (XCS) opens the possibility of simple minimally invasive surgical procedures to enable precise, long-lasting, and at-home neuromodulation with tiny implants that never contact the surface of the brain.

---

When pharmaceutical options are ineffective, under effective, or produce intolerable side effects, patients and clinicians are more frequently turning to neuromodulation for effective therapies. In the case of Parkinson’s Disease (PD) and Essential Tremor (ET), deep brain stimulation is the standard of care for treating tremor ^1^ and other symptoms ^2^. For mental health conditions like major depressive disorder (MDD), and obsessive compulsive disorder (OCD) there is a growing consensus that neurophysiologically modulating the activity in specific brain areas provides an effective therapy when drugs provide inadequate treatment ^3^. Transcranial magnetic stimulation (TMS) is one example of a method for applying this stimulation. TMS can non-invasively activate small areas of the brain surface ranging from a few millimeters to a few centimeters across using a 1-2 Tesla externally generated magnetic field ^4^, and has found success in numerous clinical studies for the treatment of neuropsychiatric conditions. Since 1998 the number of clinical trials that use TMS to address neuropsychiatric conditions has grown exponentially with a doubling time of approximately 2.5 years ^5^. Based on data from these clinical trials, the FDA has approved TMS for treatment resistant depression, and most insurance companies reimburse for multi-session in clinic therapy if patients fail traditional antidepressant therapy ^6^.There is also promising data that TMS can be used to treat obsessive compulsive disorder ^7^, PTSD ^8^, and Alzheimer’s disease ^9^. While TMS is a clinically proven therapy there are two major limitations to this therapy. First, TMS systems currently require large peak powers of approximately 3 MW ^10^, which means they are currently only approved for use in a clinic. As a result, TMS is inaccessible to patients who do not live close to a TMS facility or can’t take the time away from work or other life activities to attend daily TMS treatments. Second, session-to-session targeting can be imprecise because the stimulator must be aligned each time the patient is in the clinic. While there are other non-invasive forms for brain stimulation like transcranial direct current stimulation (tDCS) and transcranial alternating current stimulation (tACS), no at-home neural stimulation technology is in widespread use, likely because the electrical fields produced by non-invasive electrical stimulators cannot reach the field strengths needed to directly activate brain regions without also activating nerves in the scalp that produce painful side effects ^11^.

Implantable stimulators can deliver precise electrical stimulation that activates the brain continuously, but these implants require a sophisticated surgical procedure with costs and complexity that can limit patient adoption. Implantation of a chronic stimulator traditionally consists of a battery-powered implantable pulse generator (IPG) connected with wires to the stimulation site ^12–15^. When IPGs are implanted in the chest, the leads must traverse the head and neck where frequent movement results in lead migration and fracture in a reported 4% to 15% of implanted leads ^16,17^. Alternatively one can implant the IPG in the skull, which requires a patient to receive a craniotomy ^18,19^. Nevertheless, these types of devices have been successfully used to treat mood disorders making the case that long-term relief from psychiatric conditions is possible with direct electrical stimulation from implants. Electrodes placed over the dorsolateral prefrontal cortex (DLPFC) - the same target that is FDA approved for TMS therapy for depression - showed antidepressive effects in 3 of 5 patients in an early feasibility study ^12^. Deep brain stimulation (DBS) of several targets below the cortex have also shown dramatic antidepressive effects ^20^ and relief from OCD ^21^. Neuromodulation can also be achieved without an IPG by using percutaneous leads that connect to an external stimulator; however, infection from these externalized devices and the delicate wiring place limits on the patient’s lifestyle and are not commonly used for long-term neuromodulation therapies ^22,23^.

While there is a growing field of wireless battery-free stimulation technologies, these have yet to demonstrate epidural cortical stimulation in a large-animal model. Clinical battery-free neuromodulation systems have thus far only been applied in the peripheral nervous system and have not been miniaturized to the point that could be embedded within the thickness of the skull ^24–26^. Preclinical work in rodents has shown battery-free cortical stimulation ^27–29^ but the stimulation amplitudes are too small to stimulate the human brain through the dura. The only large animal, battery-free epidural cortical stimulator the authors are aware of was reported in a feline model, but the translational misalignment of only ∼3 mm makes it difficult to achieve proper alignment in our use cases in the operating room (OR) or at home ^30^.

We hypothesized that with advances in wireless power transfer (WPT) technology, we could create a cortical brain stimulator with sufficient energy and misalignment tolerance to be a practical analog to TMS, but be small enough to be implanted into a roughly 14 mm standard burr hole in the skull without ever contacting the brain surface. By implanting the device in this “minimally invasive” procedure that involves no contact with the brain or perforation of the protective dura, we aim to create an “externally powered cortical stimulation” (XCS) therapy that can be as effective as TMS and as precise and convenient as traditional battery-powered implantable stimulators.

Here we demonstrate the first millimeter-scale leadless brain stimulator in a human subject. This Digitally programmable Over-brain Therapeutic (or DOT) is approximately 9 mm across yet is capable of receiving enough energy to stimulate human and large-animal brain activity on-demand through the dura. The DOT is sealed in a glass package and maintains reliable operation in freely behaving pigs over the course of 1 month. This device could enable at-home XCS for therapeutic treatment of a variety of psychiatric disorders.

A major challenge to creating the DOT is delivering enough power to stimulate the brain from the epidural space. To meet our millimeter-size size requirements we designed the DOT to include no implanted batteries since the battery is the largest volume component of implantable neural stimulators ^31^. As a result, we needed a technology to safely deliver enough wireless power to generate the 10-15 V compliance needed for epidural brain stimulation across an impedance of ∼1 kΩ ^32^. This is comparable to spinal cord stimulation compliance ^33^, but roughly 5x the stimulation amplitude needed for vagal nerve stimulation (VNS) ^34^ and 3x the stimulation amplitude needed for deep brain stimulation (DBS) ^35^.

## Results

Recent development of magnetoelectric wireless power now makes it possible to reach the required 14.5 V stimulation compliance in millimeter sized, battery-free implants. Our lab has recently demonstrated up to 56 mW of WPT using magnetoelectric films (W. Kim, A. Tuppen, et al. *in prep)* with usable rectified voltages up to 10 V. To efficiently use this energy and enable 14.5 V compliance across 1 kΩ electrodes, we implemented a circuit using completely off-the-shelf available embedded systems components (**Fig. 1c**). Briefly, the circuit includes an efficient power rectification scheme, low-power microcontroller, uplink communication switch, and boost-converter with 15 V compliance. We then designed a custom glass enclosure that includes a stimulating and reference electrode composed of sputtered iridium oxide at the top and bottom surfaces (see methods). The completely packaged device is 9 mm by 9 mm by 11 mm.

**Fig. 1.**
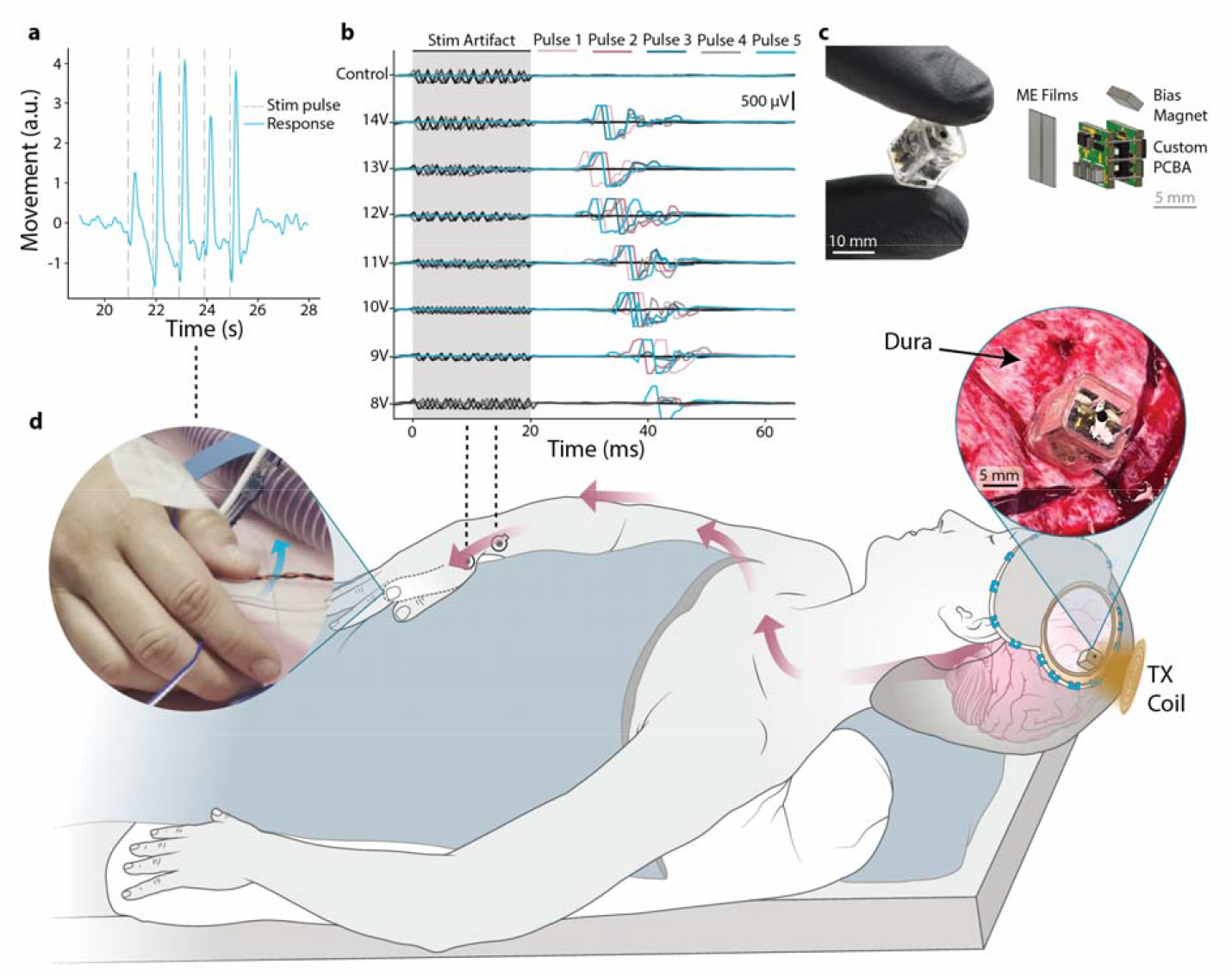
Intraoperative demonstration of epidural cortical stimulation with a millimeter-sized battery-free device. **a**, Displacement of the thumb extracted from videos during epidural cortical stimulation show movement in response to each stimulation pulse train applied by the DOT (dashed vertical lines). The DOT applied 500 Hz, 10 pulse, trains at 1 Hz frequency and 12 V biphasic amplitude. **b**, EMG traces recorded from 5 pulses each in the APB-ADM muscle groups during stimulation at varying amplitudes show that epidural cortical stimulation drives a motor response with a motor threshold around 8 V, which is the stimulation amplitude where approximately half of the pulses successfully recruit motor fibers. c, Photograph of the prototype DOT held between two gloved fingers. An illustration shows the internal components of the DOT consisting of 2 ME films, 2 panels of custom printed circuit board (PCB) connected with wires, and a bias magnet. **d**, A schematic of the intraoperative human studies, where we placed the device above the dura on the motor cortex and activated it with the transmitter coil. Stimulation resulted in contralateral thumb movement. The left inset shows a frame of the movie (**Supplementary Movie 2**) used to analyze the thumb movement and the right inset shows a photograph of the DOT placed over the dura.

To power and communicate with the DOT we fabricated a magnetic coil and driver system that transmits power and data to the implant. The digital data stream produced by the magnetic field driver programs the implant to one of 32 stimulation amplitudes between 6.75 V and 14.5 V and sets the stimulation pulse width and type of desired stimulation pulse.

### DOT demonstrates the ability to stimulate the human motor cortex from above the dura

Intraoperative testing during a neurosurgical procedure demonstrated that the DOT could stimulate a motor response when placed directly on the motor cortex with enough energy to drive a hand contraction (**Supplementary Fig. 1, Supplementary Movie 1**). In this study a patient was undergoing a procedure for tumor resection that required a craniotomy to expose the motor cortex. As part of this procedure the surgical team mapped the motor cortex using standard electrophysiological procedures to identify the region of motor cortex that, when stimulated, produced a hand contraction (see methods). We then provided the DOT and transmitter to the surgical team who placed the DOT on the identified region of the target motor cortex and placed the wireless transmitter above the DOT. After programming the DOT to produce 500 Hz, 10 pulse, trains at 1 Hz frequency and 14.5 V biphasic amplitude we observed and analyzed a video recording of 1 Hz contraction in the hand confirming that we can activate substantial regions of the motor cortex on-demand. Similar hand contractions can be generated when TMS is targeted to the motor cortex ^36^ suggesting that the volume of tissue being recruited is likely to be comparable to the volume of tissue excited by TMS.

A second intraoperative study shows that the DOT is also able to stimulate similar motor responses when placed above the dura (**Fig. 1, Supplementary Movie 2**). These data show that despite the small form factor of this millimeter-sized implant it can recruit a similarly sized region of human cortex as TMS. In this study we followed the same protocol as our first patient, but once we identified the target in the motor cortex we laid the dura back on top of the brain and placed the DOT over the dura (**Fig. 1d, right inset**).

With the dura in place, the DOT elicited visible thumb movements above 10 V stimulation amplitude (**Fig. 1d, left inset**). Analysis of video recordings with DeepLabCut analysis software ^37^ show clearly distinguishable muscle responses with each stimulation pulse (**Fig. 1a**). Electromyography (EMG) responses in the APB-ADM (Abductor pollicis brevis, Abductor digiti minimi) muscle groups were recorded at amplitudes as low as 8 V, as the stimulation intensity increased, the latency between stimulation and response decreased and the number of elicited movements increased (**Fig. 1b**). This precise localization of stimulation response demonstrates the fine precision of the DOT’s activation area. This shows that the high voltage compliance of the DOT is necessary and adequate to activate tissue in the human motor cortex from above the dura due to the increased distance and additional interposing tissue.

### Backscatter communication allows bidirectional data transfer

In addition to receiving wireless power and data, we designed the DOT to transmit data back to an external transceiver so that we can record diagnostic properties of the implant and enable future features like recording of physiological signals. To minimize size, complexity, and power demands of the implant, we use the ME material for both downlink and uplink communication ^38^. Here we implement a communication scheme where each 3.4 ms duration message has a downlink section containing 3 bits and an uplink section containing 8 bits. Stimulation parameters and other instructions can be controlled using the downlink portion of the message and status updates and current measurements can be transmitted by the device using the uplink portion of the message.

Using this data downlink we were able to wirelessly adjust simulation output and timing using the external transceiver. We designed the DOT to deliver the stimulation output at the end of the stimulation downlink command. In this way we can send instructions for the desired frequency and biphasic pulses or pulse trains can be triggered at a frequency of up to 250 Hz. For frequencies higher than this, such as the 500 Hz motor stimulation pulses, we can program the pulse train information into the device firmware and trigger it on demand with the external transceiver. **Fig. 2a** shows the timing of stimulation corresponding with the associated downlink and uplink messages. For these studies we programmed the device to output stimulation pulses between +/-6.75 V and +/-14.5 V in 250 mV increments.

**Fig. 2.**
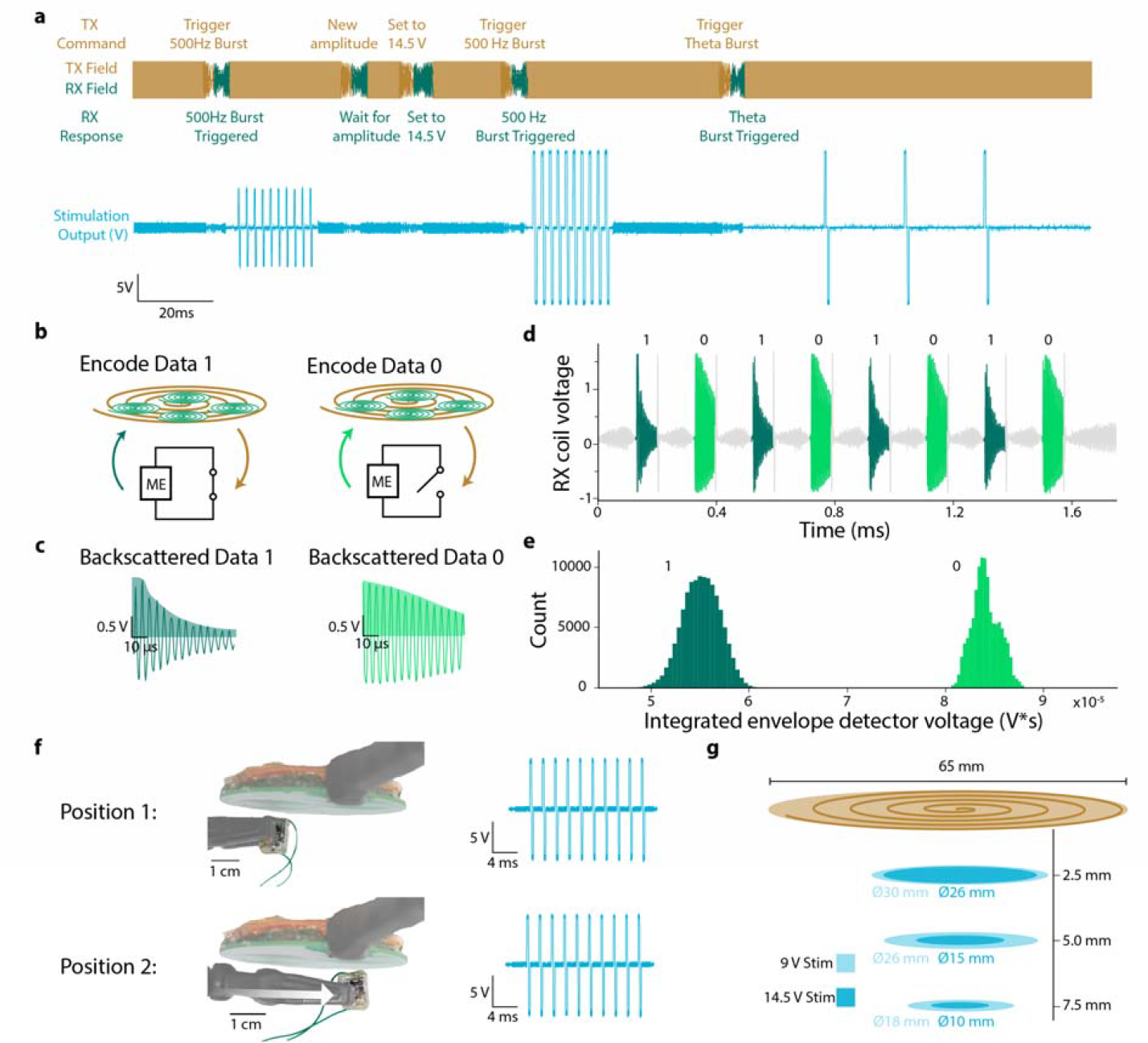
Digital bi-directional communication, and performance characteristics of the DOT. **a**, Example experimental sequence of the transceiver field and implant stimulation outputs. The transmitter field (orange) provides power during the large amplitude stimulation and charging phases, followed by reprogramming phases where digital downlink data is transmitted to the device. Device outputs are triggered by 3.4 ms duration on/off sequences in the transceiver magnetic field. We then interrogate the device settings using the passive backscatter sequence (green), that confirms the programmed settings. Here the device was programmed to have reprogrammable output voltages, stimulate with 10 pulse 500 Hz pulse trains, and stimulate with 3 pulse 50 Hz pulse trains, the changes to that stimulation pattern are written above each programming phase. The output voltage generated by the stimulator (blue) shows the different patterns of stimulation generated by a prototype DOT based on the transmitted instructions. **b**, Schematic showing how digital data is encoded by passive ME backscatter. Method of encoding bits from the implant. 1 and 0 bits are encoded by switching the nodes of the magnetoelectric film between short (data 1) and open (data 0). The films are excited by the transceiver field and the resulting ringdown field is measured from differentially paired pickup coils while the excitation field is off using a custom analog circuit. A short ringdown that corresponds to a short circuit load on the ME film represents a 1 and a long ringdown corresponding to an open circuit load represents a 0 **c**, Examples of the measured ringdown field during a 1 with a time constant of 40 µs (dark green) and 0 bit with a time constant of 91 µs (light green). Data is decoded as an integration of the positive half of the signal (shaded). **d**, A sequence of 1 and 0 bits used to calibrate the threshold for differentiating the two bits. This calibration sequence is used on demand to account for changes in implant positioning relative to the transceiver. **e**, A histogram showing the integrated voltage of the ringdown signal for bits 1 and 0 with 223888 sampled bits with the implant at a distance of 1 cm from the transceiver coils. Bits 1 and 0 are very well separated and easy to differentiate from the receiver. **f**, Experimental images showing two positions where the device is able to output biphasic 9 V, 10 pulse, 500 Hz pulse trains across a 1 kΩ resistor with a field strength of 7 mT at the coil surface and the corresponding simulation output. The distance between these two positions is over 2.5 cm, demonstrating a large translational misalignment tolerance that was important for reliable pairing in the OR. **g**, Plot showing experimentally determined results where the device is able to output biphasic 9 V or 14.5 V, 10 pulse, 500 Hz pulse trains across a 1 kΩ resistor at different distances from the transceiver coils with a field strength of 7 mT at the coil surface. At 7.5 mm distance the device is able to output 14.5 V pulse trains across a 1 cm diameter and 9 V pulse trains across a 1.8 cm diameter.

To receive real-time diagnostic information from the implant we implemented uplink communication using on/off keying of the backscattered magnetic field, which consumes almost no energy from the implant.

Briefly, we turn on the transmitter field to excite the ME film resonance mode and then turn it off to record the residual magnetic field generated as the ME resonance decays (aka “ring down”). A switch on board the implant modulates the amplitude of this reflected signal (**Fig. 2b**). If the output of the ME film is electrically connected to our stimulation circuit, the ring down will be long, but if the output of the film is electrically shorted, the ring down is much shorter (**Fig. 2c**). The resulting 0 and 1 bits are easily separable at the transceiver side with an analog circuit that integrates the envelope of the ring down signal (**Fig. 2d,e**, see methods).

The misalignment tolerance of the power and data transfer technology enables reliable wireless data and power transmission in uncontrolled environments like the OR. Good misalignment tolerance is a known quality of ME power transmission ^39^, but the exact degree of tolerance depends on the entire system configuration. To quantify the alignment tolerance for our system, intended for XCS, we adjusted the transceiver to produce a 7 mT magnetic field (within the safety limits at this frequency ^40^) at the surface of the coil, and measured the locations at which the device received enough power to produce ten 250 us per phase stimulation pulses at 500 Hz across a 1 kΩ resistance (**Fig. 2f**). When outputting 9 V biphasic stimulation pulses, the device was able to operate across a diameter of 1.8 cm (2.5 cm^2^ area) at the center of the transceiver coil at a distance of 7.5 mm from the coil (measured to the center of the top of the device). When outputting maximum compliance 14.5 V biphasic stimulation pulses the device was able to operate across a diameter of 1 cm (0.78 cm^2^ area) at a distance of 7.5 mm from the surface of the coil (**Fig. 2g**). These distances compare favorably to expected implantation depth based on reported 5.8 mm average human scalp thickness ^41^.

### Chronic study shows longevity and safety

Our intraoperative studies confirmed the DOT was powerful enough to stimulate the motor cortex through the dura, but we also wanted to confirm that our device would be able to provide this level of stimulation, safely, over time if it was chronically implanted. For this study, we used a porcine model, which is commonly used because the porcine brain anatomical structure ^42^ and dural thickness ^43^ are most consistent with human anatomy. In this preparation we tested the DOT’s ability to stimulate through intact dura and the impact on tissue response over time.

The implantation surgery took approximately 30 minutes and involved no contact with the brain (**Fig. 3a**). Briefly, we exposed the skull and drilled a 14 mm diameter burr hole over the motor cortex, exposing the dura beneath (see Methods). We then assembled the implant with a protective silicone spacer, placed it into the burr hole, and secured it with a plastic (PEEK) burr hole cover to protect the implant from external damage. A schematic of the fully implanted system and testing protocol is shown in **Fig. 3c**. This simple method was sufficient to protect and secure the implants, with none of them being mechanically damaged for the lifetime of our chronic study.

**Fig. 3.**
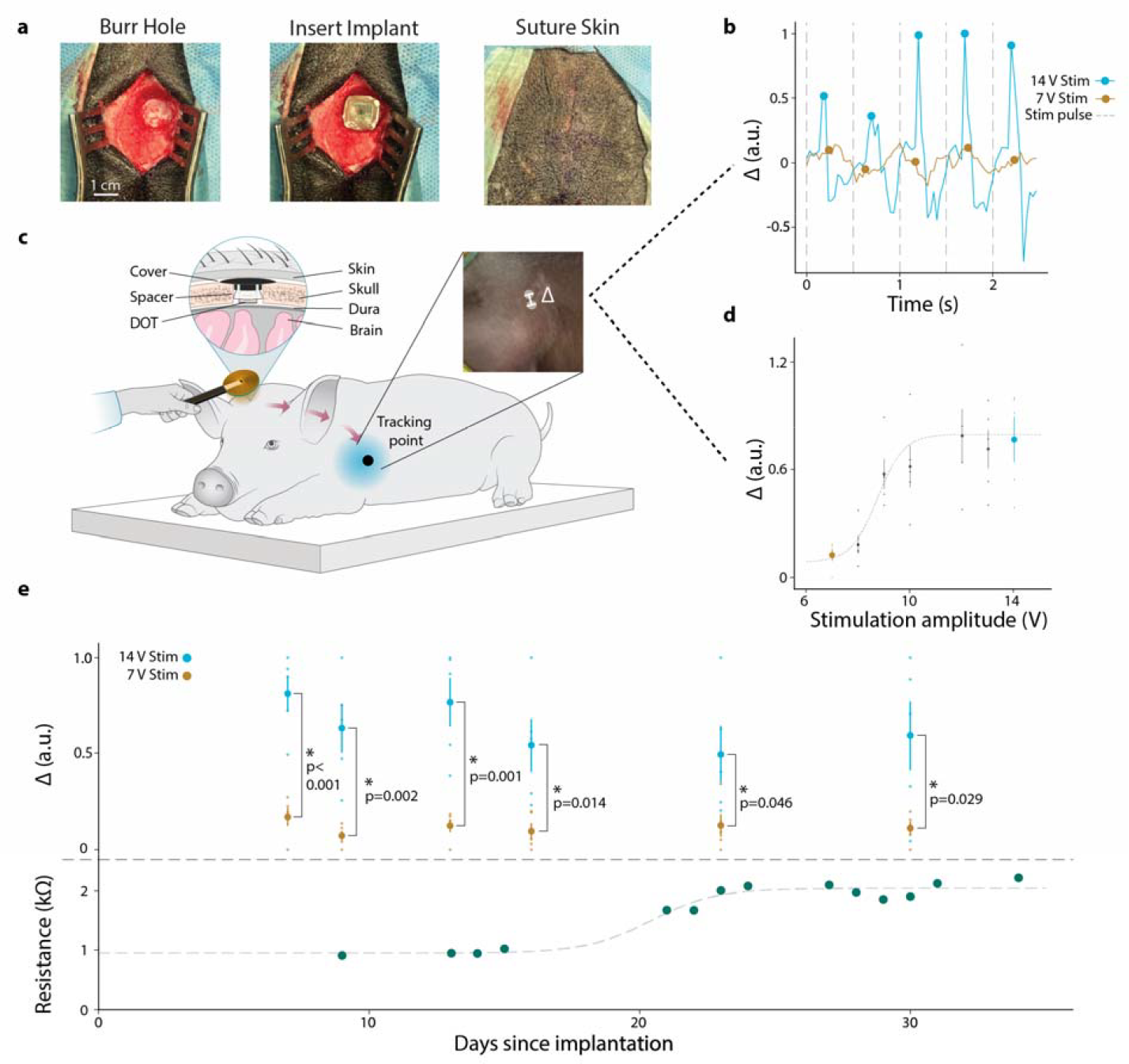
Chronic epidural cortical stimulation in a large animal model shows the DOT effectively recruits cortical activity over 30 days. **a**, Surgical photos of the DOT implantation procedure: first a 14 mm burr hole is drilled, then the device is placed in the burr hole, finally after a burr hole cover is placed over the device the skin is sutured shut. **b**, Forelimb movement in response to stimulation extracted from experimental videos using DeepLabCut. The peaks of the movement in the 350 ms following stimulation at 7 V stimulation amplitude (yellow) and 14 V stimulation amplitude (blue) are marked with circles. **c**, Schematic of device testing during chronic implantation in a porcine model. The schematic is reflected for visualization. In this study stimulation was over the left motor cortex and the movement was measured from the right forelimb. The transceiver coil is held above the implantation site while the pig is relaxed and the device is powered and programmed with the external transmitter. Left inset shows a cross section of the DOT implant above the dura and below the skin of the pig. Right inset shows an example frame from the video used to quantify forelimb movement. **d**, Mean of 5 movement peaks extracted from a video with increasing stimulation amplitude from 7 V to 14 V, error bars represent standard error. Dotted line shows a sigmoidal fit with an approximate motor threshold at 10 V. The 7 V (yellow dot) response is well below the motor threshold which we use as the noise floor and the 14 V (blue dot) response shows much higher displacement, indicative of movement in response to stimulation. **e**, (top) Mean of the movement peak amplitudes extracted from videos over the entire implantation period. Blue dots show the average movement amplitude of 5 pulses of 14 V stimulation and orange dots show the average movement amplitude of 5 samples at 7 V stimulation, error bars represent standard error. On all days tested we see significantly more motor evoked response from 14V stimulation compared to 7 V stimulation suggesting the DOT remained functional and effective at recruiting motor cortex over the entire implant period of 30 days (independent samples t-test, p<0.05). (bottom) Resistance as measured with ME backscatter communication over the

When implanted in pigs for 20-35 days above the dura over the motor cortex the DOT was able to consistently drive motor stimulation. We observed movement in response to stimulation over the entire chronic experiment in n=3 animals (**Fig. 3b,d, Supplementary Fig. 2**). We also used a sham implant in some animals to compare the pathology over time (see Methods, **Supplementary Table 1**). Movement amplitudes following stimulation were extracted using video analysis tools when possible. The results for all 30 days are shown in **Fig. 3e** (top) normalized by the minimum and maximum amplitudes from each day. We also regularly interrogated the device using our ME backscatter uplink (in n=1 animal) to record the impedance of the DOT electrodes. Between days 15 and 25 we observed an increase in resistance (**Fig. 3e**, bottom) similar to that seen with DBS electrode impedance ^44^. Between these days, study personnel noted a decrease in movement amplitude at maximum stimulation values that correlated with the increase in resistance. After 25 days the resistance stabilized at about 2.25 kΩ, which could be due to anatomical changes in the animal including thickening of the dura or tissue regrowth in the burr hole cavity. These are known foreign body responses that can occur in subdural and epidural implants, but do not present a safety concern ^45^. The fact that the DOT continued to deliver effective brain stimulation despite this increase in impedance supports its effectiveness as a chronic therapy even in the presence of foreign body responses.

When we approximated the therapeutic stimulation doses that one would receive in a TMS-like therapy, we observed no serious pathology in the stimulated brain areas suggesting that the DOT stimulation may provide a safe analog to TMS therapies. For these studies we chose to approximate the intermittent theta burst stimulation (iTBS) paradigm used by TMS to stimulate cortical areas with specifically timed pulses over the course of 3 minutes. This waveform consists of 50 Hz bursts of 3 pulses repeated at a rate of 5 Hz and is traditionally repeated for 2 seconds at 10 second intervals. Studies have shown that variations of this stimulation protocol repeated 5 days a week for 6 weeks provides effective therapy for patients with treatment resistant depression ^46,47^. The total dose received in such a therapy amounts to a total of 60-450 minutes of stimulation and a total of 12,000-90,000 pulses ^48,49^. In testing the safety of DOT stimulation we applied direct electrical stimulation of an iTBS-like waveform with 250µs per phase biphasic pulses for a total of 300 minutes (54,300 pulses). In two other animals we stimulated for a total of 170 and 200 minutes of iTBS-like stimulation. We then explanted the devices and survived the animals for 7-10 days before sacrificing the animal to analyze the brain and dura. Pathology reports of one animal post explantation, showed no difference between the brain or dura beneath the stimulator when compared to a sham implant of the same size and shape implanted in the same animal on the opposite side of the brain (**Supplementary Fig. 3**). In some animals we found minor inflammatory responses to both the active and sham implants, suggesting animal-to-animal variability in their foreign body responses, but even in the case of this foreign body response the DOT could still generate effective motor cortex stimulation suggesting that this response would not limit its ability to provide effective neuromodulation therapies.

## Discussion

Disrupting the traditional neuromodulation architecture with a millimeter sized battery-free implant could enable new paradigms for clinical therapy. For example, episodic neuromodulation that is known to be effective for TMS may one day be applied safely at home with a precisely targeted implant combined with a wearable headset that need only align approximately with the neuromodulation target. In this way, therapies like TMS that require daily visits to the clinic and precise targeting by specially trained staff can be made accessible in the home to people who live far from a clinic or who may lack the means to make such frequent trips. The simplicity, safety, and minimal management burden of XCS in this episodic use case would allow patients to avoid the maintenance checks required for implanted batteries and leads, especially rechargeable batteries that have to be periodically charged to maintain battery health. Furthermore, the XCS concept could be extended to stimulate multiple brain areas to drive specific patterns of activity or network states. For example, bi-lateral TMS over the left and right dorsolateral prefrontal cortex has shown potential as a more effective therapy for people with suicidal ideation ^50^. The XCS architecture would allow this type of multisite neuromodulation without the complexities of routing multiple lead wires as would be required for existing neuromodulation technologies.

The lack of leads or batteries greatly simplifies the surgery and eliminates many of the hazards associated with an implanted battery, but many questions and engineering challenges remain to translate this proof-of-concept DOT into a chronic human implant. Manufacturing and testing must be completed under the regulatory standards required for FDA approval including long-term safety and biocompatibility testing. Additional work is also needed to confirm that the same patterns of stimulation that are effective for TMS will also be effective for XCS. The physiological responses show that the DOT, like TMS, can recruit millimeter to centimeter sized areas of the cortex with precise timing, but the clinical equivalence in terms of therapeutic outcomes can only be confirmed once implants are approved by the FDA for chronic human studies. Nevertheless, there is promising work from implanted epidural cortical stimulators based on traditional IPG and lead technology, suggesting that electrical stimulation of the same areas targeted by TMS can produce similar clinical outcomes ^12^.

There are also additional challenges in creating a comfortable wearable transmitter. Wearable transcutaneous recharge systems have already been developed, evaluated and are in use for existing IPGs ^51^. Wearable ME transmitters have been reported for use with implants in the back ^40^, suggesting that the battery requirements and electronics can all be sufficiently miniaturized; however, a headset must be designed and tested to ensure sufficient comfort, power, and reliable pairing for frequent at-home use.

One advantage of the DOT architecture is the ability to expand the therapeutic options for neuromodulation for neuropsychiatric conditions. For example, with multiple implants that can be triggered with precise temporal patterns, one could develop more accurate ways to modulate the brain states that are believed to be associated with mood and memory ^52^. To modulate brain states with precise timing, integrating neural sensing or other sensing capabilities with these devices could allow closed-loop control of brain activity, which has been shown to improve the outcomes of TMS therapies ^53,54^. Additionally, without the requirement to travel to a clinic or sit still in a TMS chair, the frequency and duration of therapy sessions could be greatly increased, which has also been associated with improved outcomes for TRD ^48,49,55^.

Furthermore, the surgical procedure that involves no contact with the brain could increase the low eligibility and adoption rates for implantable neuromodulation which is about 5-10% for deep brain stimulation for Parkinson’s disease ^56^. Indeed simple, wireless, and battery free devices like the DOT could enable minimally invasive neurostimulation solutions for disorders that greatly reduce one’s quality of life but are not traditionally considered serious enough for IPG implantation.

With these tiny battery-free neuromodulation devices, we see a path to treat psychiatric and neurological disorders with short out-patient procedures that result in simple, long-lasting, therapies with very few side effects when compared to drugs. Indeed, with the growing trends of increasing efficacy and reduced invasiveness, neuromodulation for mental health may soon be as common as pacemakers for cardiac conditions.

## Methods

### PCBA Design

The custom PCBA is designed to harvest energy from the ME films and digitally control the output stimulation. It consists of a schottky diode bridge rectifier, storage capacitors, low-dropout regulator, boost converter (LTC3129), current sense amplifier (INA186), output switch (DG636), and Microcontroller (NXP KL15). Stimulation amplitude is controlled by setting the boost converter voltage using the digital to analog converter on-board the microcontroller. Output stimulation is switched on and off using the output switch. Charge-balanced output is maintained by stimulating with the two electrodes sequentially. First the bottom electrode stimulates with the stimulating voltage while the top electrode is connected to ground, then the top electrode stimulates while the bottom electrode is connected to ground. Output resistance is approximately measured using the current sense amplifier. The amplifier is set to monitor the supply current of the stimulation output switch. By sampling this current during stimulation output, with the knowledge of the programmed voltage, an approximation of the output resistance can be calculated. This was verified using varying stimulation voltages and load resistances to be accurate across the expected operation region.

#### Bidirectional Communication

The ME material was used for both communication from the transceiver to implant (downlink) and from the implant to transceiver (uplink). Here we implement a communication scheme where each 3.4 ms duration message has a downlink section containing 3 bits (1.8 ms duration total) and an uplink section containing 8 bits (1.6 ms duration total). We control the stimulation amplitude by sending a 3-bit amplitude command followed by a 5-bit downlink packet containing the programmed amplitude. Downlink communication is encoded in the duration of time that the magnetic field is off relative to the time that it is on. In a data 0 bit the field is off for 200 µs and on for 100 µs, in a data 1 bit the field is off for 100 µs and on for 200 µs. This is decoded on the implant by comparing the unsmoothed rectified voltage with the capacitor smoothed supply voltage (across a diode). In this way, when the field is off the rectified voltage drops below the supply voltage and when it is turned back on the rectified voltage exceeds the supply voltage. Uplink communication is encoded by the implant by switching the two nodes of the ME films between short circuit and open circuit.

After receiving a downlink message, the implant switches the uplink switch sequentially to encode the 8-bit uplink data. The transmitter pulses the field in alternating 100 µs duration excitation pulses and 100 µs ringdown periods to detect whether the encoded data is a 1 or 0. The backscattered magnetic field voltage from the ME film is received using differentially paired coils printed on a custom PCB and aligned with the transmitter coil. In this way the large magnitude excitation field is not amplified but the small amplitude backscattered field is. A custom analog front end circuit consisting of a preamplifier, variable gain amplifier, and active lowpass filter filters and amplifies the signal. An analog envelope detector smooths the output, and an onboard microcontroller (NXP LPC54606) samples this signal during the ringdown to decode the data in real time. To set the threshold between 0 and 1, a calibration signal consisting of sequential bits 01010101 is used periodically.

### DOT assembly

The DOT consists of 4 main components: the PCB assembly, two 7.5 × 3 mm ME films, a bias magnet, and 3 part glass enclosure. Briefly, ME films were manufactured by epoxying (M-Bond 43-B) PZT (Piezo Systems PZT 5H PSI-5H4E) and Metglas (2605SA1, Metglas Inc) together into a three layer laminate with Metglas on both sides of the PZT and then cutting the laminate with a femtosecond laser (One Five Origami XP, NKT Photonics). Once cut, films were tested for functionality in a 218 kHz alternating magnetic field and those with a saturation voltage above 25 V peak-to-peak were kept for use in devices. The custom PCB was designed and manufactured in separate panels by a commercial manufacturer (PCBway), then assembled by hand in the laboratory. For each device, the 2 panels were joined together using seven 3 mm lengths of uninsulated wire soldered between castellated vias on the sides of the panels. Once joined, the devices were tested for basic functionality using test firmware. Completely functional PCBAs were flashed with final firmware, then the ME films were attached in parallel to the top panel with vertically placed 0 Ω resistors and conductive epoxy. Once films were attached, the devices were tested in a magnetic field with an external bias magnet to ensure that the films provided enough power for stimulation and other functions. Then a small bias magnet was fixed in place on the assembly to allow for a wider range of motion than can be accomplished with a fixed external bias. Once operation was again verified, the test pins were cut off of the PCBA using a sanding disk attachment on a benchtop lathe.

The glass enclosure consisted of three separate parts of borosilicate glass, a rounded square tube (F&D Glass) and two caps with an electrode in the center. The caps were patterned and fabricated from a custom glass HermeS wafer (Schott) which was fabricated with hermetically sealed tungsten vias. This wafer was masked with polyimide tape to define the placement of the circular electrodes (each 1.5 mm in diameter). The wafer was then sputtered with a coating of 10 nm Ti, 100 nm Pt, 10 nm Ti, and 300 nm IrOx stack (AJA ATC Orion Sputter System) as described in previous work. ^57^ After sputtering the wafer was laser cut into rounded square caps (Universal X-660 Laser Cutter).

To assemble the final device, the bottom cap of the glass enclosure was attached with medical grade epoxy (Epo-Tek MED-301-2) to a 1 cm section of glass tube. The assembly with films was then placed inside the glass tube and stimulation output connected with conductive epoxy to the sputtered electrode on the bottom cap. This second stimulation output was connected with conductive epoxy and a ∼1 cm insulated wire to the electrode on the top cap. The top cap was then sealed with medical grade epoxy to complete the enclosure. For chronic and intraoperative studies the device was sterilized with a 12 hour Ethylene Oxide Sterilization cycle.

### DOT Characterization

Fully assembled DOT devices were tested to characterize operational performance. To measure the separability of 0 and 1 data bits, a fully assembled implant was tested with the calibration sequence. The implant was placed 1 cm above the paired transmitter and receiver coils and powered with a 7 mT magnetic field. The transceiver was programmed to send, receive, and print raw output values for 8-bit calibration signals at a rate of 50 Hz. 223888 bits were sampled and plotted with a histogram to show separability. No data points were discarded as outliers.

To measure the alignment tolerance of the system for WPT, wires were connected to the stimulation electrodes of a fully assembled implant, connected across a 1 kΩ resistor and monitored with a 1 MΩ impedance oscilloscope. The transceiver was adjusted to produce a 7 mT magnetic field at the surface. At each height the device was repeatedly triggered to output stimulation at the desired voltage and moved along a linear axis from the center of the coil until the last pulse of the 10-pulse train started to drop, this was marked as the operational region. Rotational symmetry is assumed based on the symmetric geometry of the transceiver system.

### Intraoperative human testing

The studies took place during tumor resection surgeries at Baylor St Luke’s Medical Center under IRB approval (Protocol H-50885) and complied with all relevant ethical regulations. We obtained informed consent from study participants.

Prior to using the DOT, the patient was placed under anesthesia and had the surgical site prepped and the motor cortex exposed via a craniotomy. The location of the primary motor cortex was determined by locating the central sulcus with median nerve stimulation and ECoG recording to determine the location where phase reversal occurred. The specific cortical targets within the primary motor cortex to activate with the DOT were determined from motor mapping with probe stimulator and EMG monitoring using a Cadwell IOMAX system by a neuromonitoring technician on site. In the first intraoperative study the DOT was placed on the right motor cortex, activating muscles in the left hand, while in the second patient, the DOT was placed on the left motor cortex, activating muscles in the right hand.

The physician was verbally guided as to how to place the device above the brain target, and used gelfoam to secure it on the cortical surface. In order to make electrical contact between the top electrode and the conductive body tissue (as we would expect in an actual implantation scheme), the surgeon placed a piece of wet gauze over the top of the device to the surface of the dura. The surgeon was instructed on how to place the transmitter coil by study personnel within the operating room. Study personnel powered and commanded the coil to power the implant via a computer and turned on the stimulation at the amplitude analogous to the amplitude used with the Cadwell IOMAX system and elicited a motor response.

Video recordings were taken of the patient’s hand during stimulation. Stimulation timing was announced verbally during the operation. In post-analysis, the audio cues were used to find the approximate timing of stimulation pulses. The videos were then analyzed with DeepLabCut by marking a stationary point in the scene and features on the hand and fingers. The distance between the stationary point and the hand was measured over time and the signal bandpass filtered between 0.5 and 3 Hz since the response to stimulation was at 1 Hz.

### In vivo porcine model

All procedures were approved by the University of Texas Health Science Center at Houston Animal Welfare Committee (Protocol AWC-22-0023) and complied with legal and ethical standards regarding the use of vertebrate animals in research and teaching at UTHealth.

The results presented in this work contain results from four cohorts of pigs (for a total of n=8 animals and n=10 implants, see Supplemental Table 1). The laboratory animals used for the porcine work reported here used a total of n = 8 domestic swine of two different species, n= 4 Duroc and n= 4 Hampshire. All animals weighed 20-40kg when implanted. The first two cohorts provided feasibility insight into the surgical procedure, animal handling, and electrode design. In these two cohorts, all implants were functional when implanted and the animals followed the same general chronic procedure outlined here, but no chronic stimulation was applied. The outcome of these two early cohorts resulted in changes to implantation procedure (including a burr hole cover) and changes from a bipolar (two electrodes on the bottom of the implant) to a pseudo-monopolar electrode (electrodes on the top and bottom of the implant) design for more effective tissue stimulation ^58^. All of these changes were implemented for cohorts 3 and 4. In the third cohort one sham animal received no stimulation, while the other received stimulation. In the fourth cohort each animal was implanted with two implants-one active implant that provided stimulation to the left motor cortex and one sham implant over the right motor cortex.

Each study took place in six distinct parts: (1) Pre-implantation, (2) Surgical implantation, (3) Daily testing, (4) Surgical explantation, (5) Terminal procedure and euthanasia, and (6) tissue harvest.

#### 1) Pre-Implantation Conditioning / Acclimation

Animals were received at the facility, were housed in pairs in a shared pen, and allowed to acclimate to their surroundings, including study personnel over the course of several weeks. For 1-2 hours a day, study personnel sat in the animal cage and calmly encouraged the animals to approach, lay down, and allow personnel to touch their head, face, and ears with their hands and with a testing coil. By the time of implantation, both animals were comfortable with this behavior and behaved normally around study personnel. This was performed based on previous study experiences allowing daily testing of the devices with minimum interruption to the animals’ daily activities.

#### 2) Surgical implantation

Surgical implantation began with facility veterinary staff preparing the animals on the morning of the implantation. Study personnel arrived and completed surgical implantation of the device, in approximately 30-60 minutes per animal, depending on whether any intraoperative testing was performed. ECG, respiration rate, ETCO2, SpO2, rectal temperature, and jaw tone (to assess depth) were all continuously monitored during the implant and explant procedures.

The implant procedure involved placing the DOT device flush with the skull and atop the dura via a typical neurosurgical burr hole procedure. Briefly, hair was shaved and a small incision was made above the motor cortex, midline and parallel to the eyes, ∼4 cm. A surgical window of 4 by 5 cm was cleaned and prepared for cranial burr access. Using a Medtronic Midas Rex drill with a standard cranial burr bit, a cylindrical burr hole of approximately 14 mm outer diameter was drilled down to the dura. The dura was not punctured.

During this procedure in the third cohort tested one animal was first anesthetized with isoflurane and then weaned onto ketamine. Previous work has shown that any lingering isoflurane will block the motor response that can be generated from epidural cortical stimulation ^59^. At least 10 minutes of ketamine with no isoflurane was needed to properly engage motor response. In some cases as the animal began to wake up during ketamine anesthesia, isoflurane had to be reengaged, leading to a careful interplay between the two before achieving a motor response.

In this third cohort, animal intraoperative epidural stimulation using a Cadwell IOMAX and disposable monopolar stimulator probe (Neuroradium, Inc.) verified adequate targeting above the motor cortex and, further, sufficient amplitude of stimulation confirmed the expected motor response. In addition, disposable subdural needle electrodes (Neuroradium, Inc.) were placed on the forelimb of porcine subjects to record electromyographic (EMG) activity. Data were visualized and synchronized in real time with stimulation onset to confirm motor activation thresholds and subsequent compound muscle action potentials (CMAPs). Offline analysis of these recorded files was also completed.

Once targeting above the motor cortex was confirmed, the DOT and accessories were assembled and implanted in the burr hole space (**Supplementary Fig. 2**). Prior to closing the surgical site, the DOT was driven to an electrical stimulation output sufficient to elicit a similar motor output to that observed with the Cadwell IOMAX system. Lastly, the surgical site was sutured closed and the animal went into recovery.

In the fourth cohort, no intraoperative testing results were used for targeting or stimulation verification. The DOT was simply implanted in the same area used in the previous cohort.

#### 3) Daily Testing

While animals were approved to test the day following surgery, study staff waited two days until daily testing. During these two days, staff regularly applied pain reducing medications and cleaned the incisions. Extensive monitoring occurred for 5-7 days taking note of attitude, incision health, and appetite. Study personnel were allowed to enter the pen and spent time feeding and calming the animals until they laid down for rest. Once resting, the external magnetic field driver was introduced to the pen and the therapy workflow was carried out.

Therapy workflow included (1) running a motor stimulation waveform paradigm and observing any motor output, (2) running a backscatter technique to assess bidirectional communication or measure impedance, and (3) running a therapy paradigm analogous to iTBS stimulation.

Running stimulation paradigms involved introducing the external magnetic field driver above the implant site and running a downlink communication command to the implant. During motor stimulation, if successful, a regular non-voluntary motor response was observed in the right shoulder or forelimb of the animal under test. Videos of the motor response with verbal cues when stimulation was applied were recorded when possible for later analysis.

The backscatter calibration technique was pre-programmed to execute an 8 bit command code between the implant and receiving coil. A passed calibration is one in which the expected signal (01010101) is correctly received from the implant. This calibration sets the threshold between 0 and 1 for other communication messages.

### Video analysis

In post-analysis, the audio cues were used to find the approximate timing of stimulation pulses. The videos were then analyzed with DeepLabCut by marking a stationary point in the scene and features on the area of the pig that was moving during stimulation. The distance between the stationary point and the moving point was measured over time and the signal highpass filtered above 1 Hz since the response to stimulation was at 2 Hz. Peaks were then identified in the response within a 350 ms period following the application of each stimulation pulse (5 pulses for each trial). The peak values were extracted across each stimulation level applied. Because the angle and distance of the recorded video changed each day, the relative amplitudes of movement are difficult to compare, so the resulting values are normalized to a 0 - 1 scale based on the minimum and maximum peaks recorded that day. Statistical significance between 7 V stimulation and 14 V stimulation is evaluated with an independent samples t-test (p < 0.05).

### Impedance measurement and evaluation

Output impedance is measured using the onboard circuitry during stimulation (see Methods, PCBA design). This data was transmitted via uplink during chronic testing, with each impedance measurement preceded by a calibration message to ensure proper alignment and calculate the threshold between 0 and 1 data. The transmitter was programmed to trigger the device to apply a 500 Hz stimulation burst at 7, 8, and 9 V biphasic amplitudes sequentially and measure the impedance at each level. The uplink data was logged in a text file with the envelope detector values sampled during each bit of each message. In post-analysis, measurement sequences with successful calibration at each of 7, 8, and 9 V were extracted, and the average impedance measured across these 3 values is reported as the impedance value.

#### 4) Surgical explantation

Animals were prepared similarly to implantation by veterinary staff. Before removing the device in the third cohort, a similar procedure was completed where stimulation was driven by the DOT and EMG recordings were captured via the Cadwell IOMAX system with subsequent motor output. In order to achieve this the anesthesia was tuned as explained in the implant procedure.

A similar 4×5 cm window was prepared to gain access to the previously implanted device. Using typical neurosurgical instrumentation, the devices were explanted, the incision cleaned, closed using a stapler, and the animals went into recovery. Recovery consisted of the same protocol done following implantation.

#### 5) Terminal Procedure and Euthanasia

Animals were prepared for sacrifice approximately one week after explantation. Pigs were put under heavy sedation and then sacrificed using a combination of Telazol, Glycopyrrolate, and Somnasol.

#### 6) Tissue Harvesting and Histology

After sacrifice, the brains were removed for histological examination. The brain specimens were subjected to 10% buffered formalin solution for a minimum duration of seven days. Subsequently, a meticulous brain dissection procedure was carried out to isolate the pertinent brain regions for detailed microscopic examination. For histological assessment, formalin-fixed paraffin-embedded sections with a thickness of 5 um were prepared. Hematoxylin-eosin staining and immunohistochemistry staining techniques were then applied to these sections. Specifically, ionizing calcium adapter binding molecule 1 (Iba1) and glial fibrillary acidic protein (GFAP) were employed as markers to discern and evaluate microglial and astrocyte reactivity and activation, respectively.

## Supporting information

Video of motor response to epidural XCS stimulation

Video of motor response to subdural XCS stimulation

## Reporting Summary

Further information on research design is available in the Nature Research Reporting Summary linked to this article.

## Data availability

The main data supporting the results in this study are available within the paper and its Supplementary Information. The raw data are available from the corresponding authors on reasonable request.

## Acknowledgements

The authors would like to thank Rice SEA for the use of and training on equipment, Prof. Kaiyuan Yang, Zhanghao Yu, and Matthew Parker for their helpful discussion in system design, the UTHealth Veterinary Staff for their care of the pigs used in this study, Scott Crosby for his assistance recording and interpreting electrophysiological data, Baylor St. Luke’s Surgical Staff for their care in the operating room, Steve Goetz for his helpful review of the manuscript, and Elena Kakoshina for illustrations and design of Figures 1 and 3. Supplementary Figures 1a, b and e were created using Biorender.com.

## Author contributions

JTR and AS conceptualized and designed the DOT. JW, AS, and WT assembled and tested the DOT. JW designed the DOT PCBA. FA and JW designed and tested the backscatter communication protocol and associated circuitry. SaAS, JW, and AS performed the intraoperative patient testing. SaAS and SuAS performed the porcine model surgeries. AS, WT, and JW performed the chronic large-animal testing. JW and AS analyzed the intraoperative patient and large-animal data. CT performed and analyzed the histology of porcine tissue. JW, AS, and JTR prepared the manuscript with input from all authors.

## Additional Information

