## Supplementary Information for "Millimeter-sized battery-free epidural cortical stimulators"

**Supplementary Information (SI)**

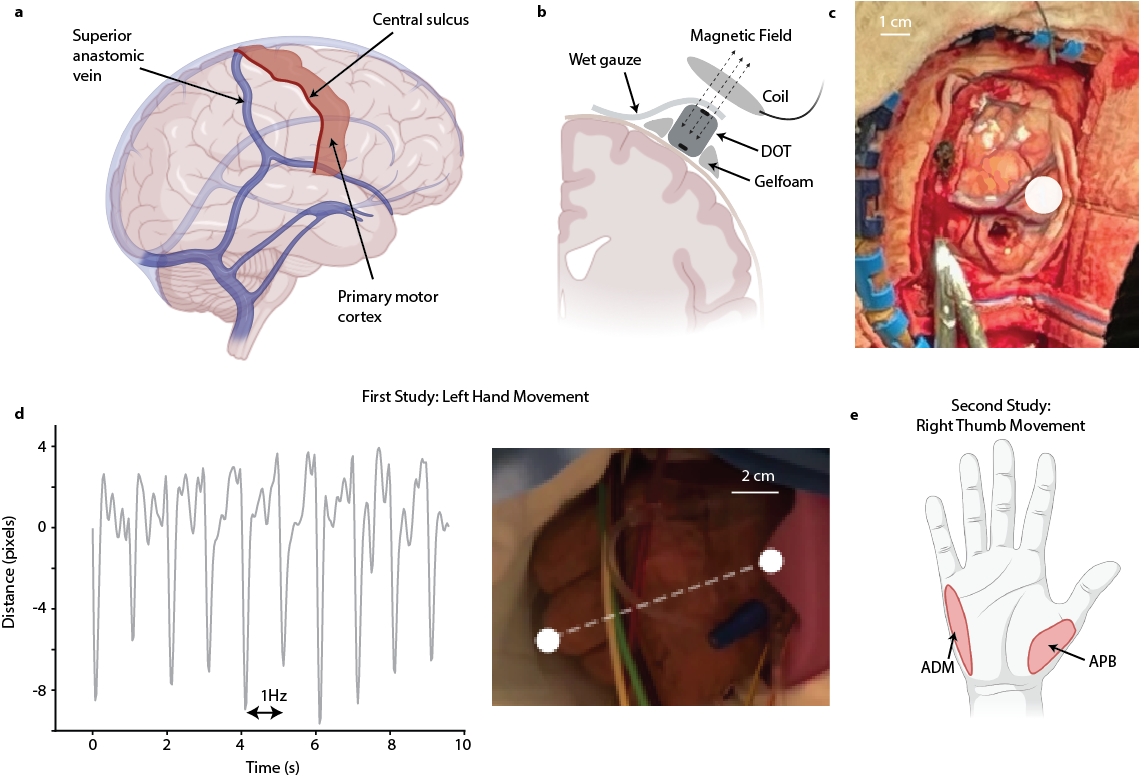

**Supplementary Figure 1 | Interoperative human supplemental schematics and data. a**, Anatomy of the right brain hemisphere showing relevant anatomical markers and the location of the primary motor cortex where stimulation was applied. **b**, Schematic showing the setup of the DOT on the dura/cortical surface secured in place with gelfoam and making electrical contact to the top electrode with gauze. **c**, Photograph of the surgical site for the first intraoperative patient with the location of the DOT on the motor cortex denoted with a white circle. **d**, Results from the first intraoperative human study show movement of the fist clenching in the left hand. Video analysis shows this movement is consistent with the 1 Hz stimulation applied by the DOT. **e**, Schematic showing the location of the muscles in the right hand where results from the second intraoperative study (Main text, Fig. 1) recorded EMG activity (ADM-APB electrode channels). The location of these muscles is consistent with the thumb movement recorded visually.

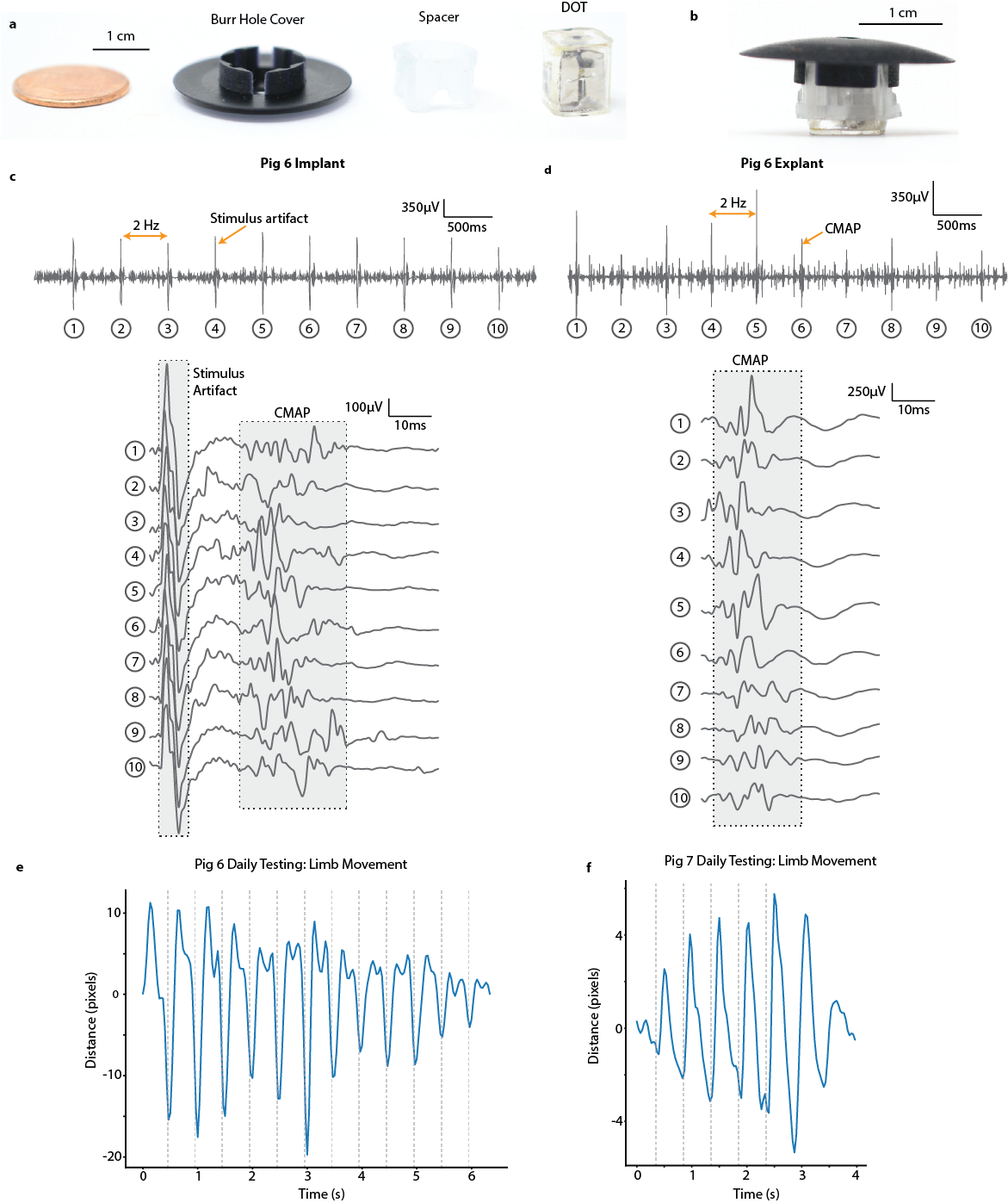

**Supplementary Figure 2 | Additional in vivo porcine results. a**, The parts of the implant assembly-including the burr hole cover to protect the implant from impact, the spacer to fill in the rest of the 14 mm burr hole and the DOT-unassembled with a 1c coin and **b**, assembled for implantation. **c**, EMG recordings of muscle activity in the right leg of pig 6 activated by DOT stimulation of the motor cortex taken during implantation, and **d**, explantation verifying chronic functionality. **e**, Video analysis example of motor movement in the right side in freely behaving animals in Pig 6 and **f**, Pig 7 demonstrating chronic stimulation functionality across multiple animals.

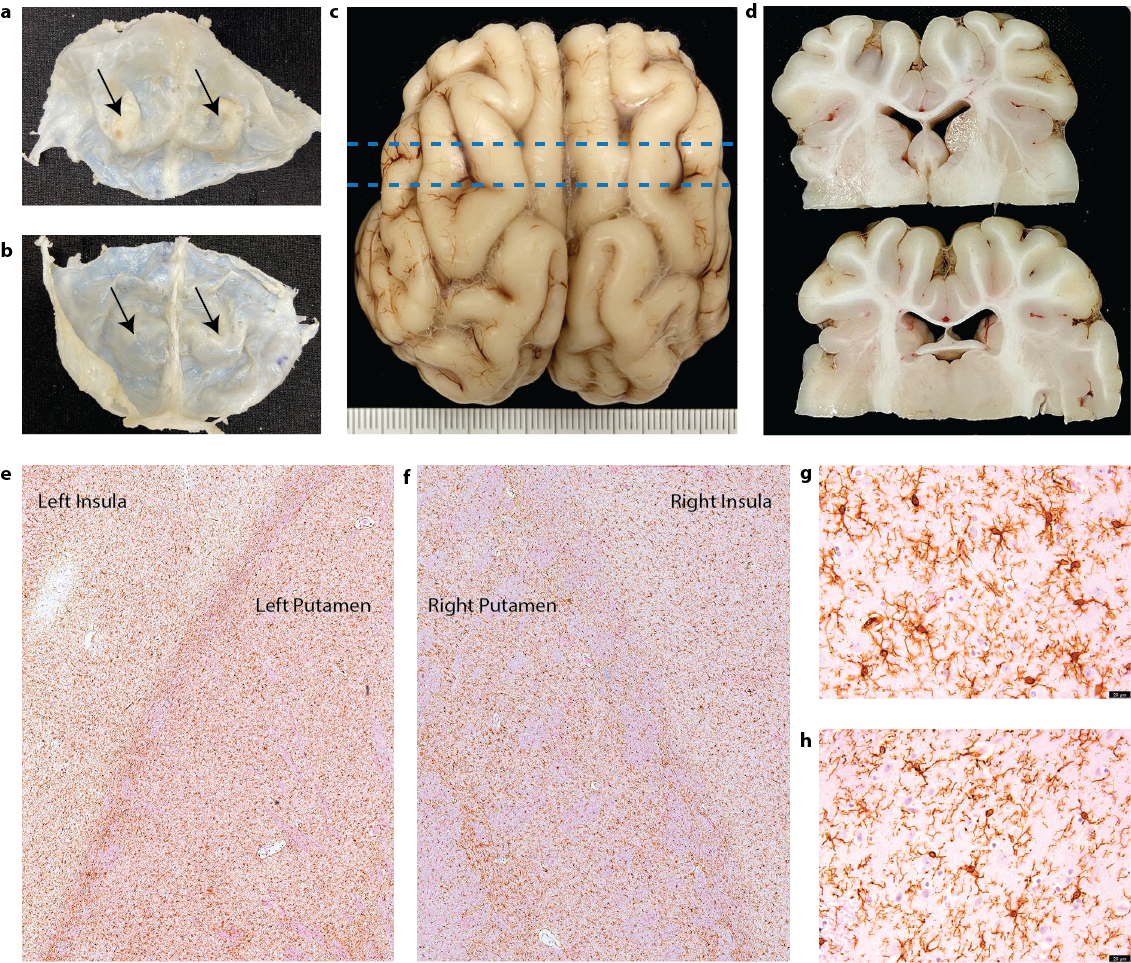

**Supplementary Figure 3 | Porcine brain histology results show no signs of pathology or damage to brain tissue in bilateral implantation comparison. a,** Top and **b,** bottom of the dura show thickening on the top of the dura where the implant (left) and sham (right) were located (denoted with arrows) **c,** Convexity view of the brain shows no gross cortical damage, blue lines indicate the locations of the views of the slice shown in **d,** which again shows no tissue damage. Iba1 immunostaining for microglia shows expression on both the left (**e, g**) and right (**f, h**) sides with slightly increased number of microglia in the left side which can be expected as a result of stimulation.

| **Cohort** | **Pig/Implant Side** | **Minutes of stim** | **Days implanted** | **Dura thickening** | **Focal brain tissue loss** | **White matter microglia** | **Infection at surgical site** | **Implant damaged** |
| --- | --- | --- | --- | --- | --- | --- | --- | --- |
| 1 | Pig 1/L | Sham | 21 | Y | N | N | N | N |
|  | Pig 2/L | Sham | 21 | X | X | X | N | Y* |
| 2 | Pig 3/L | Sham | 18 | X | X | X | N | N |
|  | Pig 4/L | Sham | 18 | X | X | X | N | N |
| 3 | Pig 5/L | Sham | 20 | Y | Y | Y | N | Y** |
|  | Pig 6/L | 170 | 20 | X | Y | Y | N | N |
| 4 | Pig 7/R | Sham | 35 | Y | N | Y | N | N |
|  | Pig 7/L | 200 | 35 | Y | N | Y | Y | N |
|  | **Pig 8/R** | **Sham** | **35** | **Y** | **N** | **Y** | **N** | **N** |
|  | **Pig 8/L** | **300** | **35** | **Y** | **N** | **Y** | **N** | **N** |

Y = Yes, N = No, X = No results *No burr hole cover – broken glass, **Epoxy failure

**Supplementary Table 1 | Porcine Cohort Summary.** Summary of the four porcine cohorts used in this work indicating the implant location, how much stimulation was applied, histology notes, and any adverse events. Bold text denotes the animal used in the brain analysis in supplementary figure 3 and main text figure 3.

**Supplementary Movie 1 | Left hand movement in response to subdural XCS** In the first interoperative study the DOT placed directly on the right motor cortex and activated at 1Hz activated left hand clenching at 1Hz.

**Supplementary Movie 2 | Right hand movement in response to epidural XCS** In the second interoperative study the DOT placed on the dura above the left motor cortex and activated at 1Hz activated thumb movement in the right hand at 1Hz.
